# Acceptability and Feasibility of Genetic Testing to Assess Risk of HIV-Associated Neurocognitive Impairment among Thai Adolescents and Young Adults

**DOI:** 10.1101/2022.04.28.22274059

**Authors:** Anthony F. Santoro, Christopher M. Ferraris, Maral Aghvinian, Linda Aurpibul, Dennis Kolson, Reuben N. Robbins

**Affiliations:** HIV Center for Clinical and Behavioral Studies, Columbia University and New York State Psychiatric Institute, New York, NY, USA; Department of Psychology, Fordham University, New York City, NY, USA; Research Institute for Health Sciences, Chiang Mai University, Chiang Mai, Thailand; Department of Neurology, Perelman School of Medicine, University of Pennsylvania, Philadelphia, PA, USA

**Keywords:** HIV-associated neurocognitive impairment, genetic testing, acceptability, perinatally-acquired HIV, youth living with HIV

## Abstract

Host genetic factors may modify the risk of developing HIV-associated neurocognitive impairment (HIV-NCI), and genetic research has the potential to inform novel treatments for HIV-NCI. However, there is a need to better understand the acceptability of genetic testing among distinct populations of people living with HIV at increased risk for HIV-NCI, such as young people living with perinatally-acquired HIV (PHIV) in low- and middle-income countries (LMICs), to gauge the feasibility of genetic research within these populations. This pilot study evaluated the acceptability and feasibility of genetic testing to assess risk of future neurocognitive problems in fifty Thai adolescents and young adults (12-24 years; *M*_*age*_=19.16; 52% female) with PHIV and demographically similar HIV-negative controls. Participants completed a survey assessing acceptability of and concerns about genetic testing and were asked to provide blood samples for genetic testing. Descriptive statistics and blood draw completion rates were produced and calculated. Reported concerns about genetic testing were grouped thematically and tallied. Independent *t* tests and chi-squares explored demographic differences between participants who reported concerns and peers. Results indicated 46 participants (92%) rated genetic testing as “acceptable” or “completely acceptable.” Eight participants (16%) reported concerns about genetic testing. The most common concerns were related to genetic information being shared or misused. Compared to peers, participants who reported concerns had more years of education and were more likely to have post-secondary schooling. Regarding completion rates, 49 participants (98%) agreed to genetic testing and provided blood samples. Overall, results support the acceptability and feasibility of incorporating genetic testing into research investigating HIV-NCI among adolescents and young adults in Thailand. Findings provide important considerations for planning future genetic studies among young people in Thailand and perhaps other LMICs.

## Introduction

Globally, approximately 5 million young people (15-25 years) are living with HIV, many with perinatally-acquired HIV and most are residing in low- and middle-income countries (LMICs).^1^ Considering youth with perinatally-acquired HIV (PHIV) are exposed to HIV in their central nervous systems for the entirety of neurodevelopment, young people with PHIV may be at particular risk for developing HIV-associated neurocognitive impairment (HIV-NCI).^2^ HIV-NCI is one of the more common non-communicable complications of HIV infection and is associated with a host of poor disease and functional outcomes, including suboptimal medication adherence, psychiatric symptoms, and poor academic performance.^2-4^
Although the causes of HIV-NCI in the context of antiretroviral therapy (ART) and viral suppression remain unclear, a growing body of research suggests that host genetic factors may underlie risk of developing HIV-NCI in adults with HIV.^5-8^ For example, evidence suggests that a common genetic polymorphism (dinucleotide (GT)*n* repeat length variation) in the promoter region of the heme oxygenase-1 (HO-1) gene, which regulates the expression of cytoprotective enzyme heme-oxygenese-1, may moderate HIV-NCI risk.^6,7^ Specifically, in adults living with HIV, shorter HO-1 (GT)*n* repeat length alleles have been associated with increased expression of HO-1, reduced neuroinflammation, and decreased rates of HIV-NCI.^6,7^ Separately, apolipoprotein E4 (APOE) isoforms have also been investigated in relation to HIV-NCI risk, with evidence suggesting APOE ε4 alleles associate with cognitive decline and worse memory performance in older adults living with HIV.^8,9^

To date, most of the HIV-NCI research investigating genetic risk factors has focused on behaviorally-infected, middle-aged adults, with genetic research remaining limited among people living with HIV in LMICs, where HIV burden is greatest,^1^ and among people with PHIV, who are at increased risk for HIV-NCI.^2^ There are barriers to implementing and scaling up genetic testing in LMICs, including large costs of developing testing programs and a lack of healthcare providers (e.g., genetic counselors) to provide these services.^10,11^ However, genetic testing is becoming cheaper, and genetic testing services are quickly evolving in LMICs, primarily through research efforts and international collaborations.^10,11^

With the advent of more widespread adoption of genetic testing in research and clinical services, there is a timely need to examine the acceptability of genetic testing among distinct populations of people living in LMICs to appropriately gauge the feasibility of scaling up these services for specific populations of people living with HIV. A recent systematic review characterized several ethical, social, and cultural considerations (e.g., family values, religious laws, local customs) that could serve as barriers to the uptake of genetic testing and services in LMICs, highlighting that the acceptability of genetic testing within these regions may be highly dependent upon social influences and stigma related to having a genetic condition.^11^ As external barriers to genetic testing are increasingly reduced and genetic technology becomes more widely available, understanding patient-specific factors related to the acceptability of using genetic testing and services will become critical.^11^

In addition to better understanding the acceptability of genetic testing among people living in LMICs, there is a need for more genetic research focused on children, adolescents, and young adults.^12^ Young people are underrepresented in the genetic literature, and genetic variation in adolescents and young adults remains highly understudied, hindering the development of novel treatment approaches for youth populations.^12^ In accord, most research on genetic risk factors of HIV-NCI have focused on adults, obscuring our understanding of genetic risk factors of HIV-NCI among young people living with HIV and precluding our ability to develop tailored interventions for these young people.

The purpose of the current pilot study was to inform future research investigating the genetic underpinnings of HIV-NCI among young people living with HIV in Thailand. Specifically, we evaluated the acceptability and feasibility of genetic testing to assess risk of neurocognitive problems in a pilot sample of Thai adolescents and young adults with PHIV and demographically similar HIV-negative peers. Likewise, we assessed concerns related to genetic testing and examined the demographics (e.g., HIV status, age, education) of participants who reported concerns to further inform future studies, as well as the development of related clinical services for these youth. This pilot study was among the first to support the acceptability and feasibility of research investigating the genetic risk factors of HIV-NCI among young people living with HIV in an LMIC.

## Materials and Methods

### Design and setting

This study collected data on a pilot sample of Thai adolescents and young adults (18-24 years) with PHIV and demographically similar HIV-negative controls. Study procedures and data collection took place at the Research Institute for Health Sciences, Chiang Mai University (RIHES-CMU), Chiang Mai, Thailand.

### Participants

Fifty Thai participants (25 PHIV; 25 controls) were recruited from an ongoing parent study at RIHES-CMU examining neurocognitive functioning among Thai youth. The parent study’s inclusion criteria were a) 13-24 years old; b) perinatally-acquired HIV; c) currently on an ART regimen. PHIV participants who completed the parent study within the last year were recruited to participate in this pilot study during regularly scheduled ART clinic visits. Demographically similar (i.e., age, sex) HIV-negative participants were recruited from the local community at the time of their parent study visit.

### Procedures

This study was conducted with ethical approval from the Institutional Review Board at the New York State Psychiatric Institute and received a Certification of Ethical Clearance from the Human Experimentation Committee at RIHES-CMU. Informed consent and assent, for participants under 18 years old, was obtained from all participants. For participants under 18 years old, informed consent was also obtained from participants’ parents/guardians. Common among genetic studies, during the informed consent/assent process, study staff provided information to participants about genetic testing and related risks and protections. Participants were informed that although blood samples and genetic data would be deidentified, genetic information is unique to each person and that the risk of being identified based on this information may change in the future as new methods of tracing genetic information are developed. Participants were also informed about the possibility that health or genetic information could be misused by third parties (e.g., employers, insurance companies) and were assured that information and samples that could be used to identify them would not be released without their permission, unless otherwise required by the law. Following consent/assent procedures, participants completed survey items assessing acceptability of genetic testing and were asked to provide a blood sample for genetic testing. For participants who agreed to the blood draw, 2ml of whole blood was collected via venipuncture. Participants who declined the blood draw were asked their reason for declining, and study staff completed a form documenting the participant’s verbatim response. Participants were compensated for their time, irrespective of whether they provided blood samples. Collected blood samples were stored onsite in a -80C freezer at RIHES-CMU and cold-chain shipped in one batch to the United States (US) for processing. Study materials (e.g., consent/assent forms) were translated into Thai, back-translated into English, and modified, as needed.

### Measures

#### Acceptability

Adopted from previous research assessing the acceptability of genetic screening^13^, participants were asked: “In your opinion, how acceptable is genetic testing to screen for risk of cognitive problems in the future?” Participants recorded their response on a 5-point Likert-type scale (1 = *Completely unacceptable*; 2 = *Unacceptable*; 3 = *Unsure*; 4 = *Acceptable*; 5 = *Completely acceptable*). An average rating of 4.00 was set as an a priori benchmark of acceptability.

#### Concerns

Participants were also asked one open-ended question: “Do you have any concerns about genetic testing?” Participants recorded responses in Thai on the response form. Text responses were translated into English by study staff. A dichotomous variable was created to codify participants who reported concerns and participants who did not report concerns, coded 1 and 0, respectively.

#### Feasibility

To assess feasibility, we documented the number of participants who were approached to participate in this pilot study and the number of participants who completed blood draws for genetic testing. We calculated the completion rate of blood draws for genetic testing ([# of completed blood draws / # of participants approached] x 100). An a priori benchmark of 80% was set to indicate feasibility. We also calculated the percent of collected samples that were successfully shipped and arrived intact at the US-based laboratory for processing ([# of intact samples received by laboratory / # of samples collected] x 100).

#### Demographics

The following demographics were collected: HIV status, age, gender identity, education, and employment/student status.

### Analytic Plan

Descriptive statistics and measures of central tendency were produced to summarize demographic and acceptability data for the total sample and for PHIV and control participants, separately. Blood draw completion rates were also calculated for the total sample and for PHIV and control participants, separately. As exploratory analyses, independent *t* tests and chi-square tests were conducted to assess significant group differences between: a) PHIV and control participants and b) participants who reported concerns about genetic testing and participants who did not report concerns. Text responses to the open-ended question asking about participants’ concerns about genetic testing were translated into English, organized in a table, grouped thematically, and tallied.

## Results

### Demographics of study participants

On average, participants were 19.16 years old (*SD* = 3.09), with 30% of participants between 12-17 years old and 70% between 18-24 years old. About half (52%) of the sample identified as female. On average, participants had 11.40 years of education, and the majority (66%) was currently studying in school, college, or university; 34% reported working full-time. Regarding differences by HIV status, a significant difference was found for years of education, *t* (48) = 2.12, *p* = .04, *d* = .60, with control participants reporting more years of education (*M* = 12.24 education years, *SD* = 2.86) compared to PHIV participants (*M* = 10.56 education years, *SD* = 2.74). That said, a similar proportion of PHIV participants (48%) and controls (56%) reported at least some post-secondary education, attending/completing vocational school or college/university. Differences by HIV status were also found for student/employment status, *X*^2^ (1, *N* = 50) = 7.22, *p* = .007, with a greater proportion of PHIV participants (52%) reporting full-time employment compared to controls (16%). Conversely, a greater proportion of control participants (84%) were currently enrolled in school compared to PHIV participants (48%). PHIV and control participants did not significantly differ in age or gender identity.

### Acceptability

Table 1 presents acceptability data. On average, participants rated genetic testing as highly acceptable (*M* = 4.23, *SD* = 0.69), with 46 participants (92%) rating the use of genetic testing to screen for risk of neurocognitive problems as either “acceptable” or “completely acceptable”. No participant rated genetic testing as “completely unacceptable”. Acceptability ratings did not significantly differ between PHIV (*M* = 4.20, *SD* = 0.71) and control (*M* = 4.48, *SD* = 0.65) participants.

**Table 1.** Acceptability of Genetic Testing, Concerns, and Blood Draw Completion Rates

|  | **PHIV**<br>**(*n* = 25)** | **Controls**<br>**(*n*=25)** | **Total**<br>**(*N*=50)** |
| --- | --- | --- | --- |
|  | ***M* (*SD*)**<br>**[*Range*]** | ***M* (*SD*)**<br>**[*Range*]** | ***M* (*SD*)**<br>**[*Range*]** |
| **In your opinion, how acceptable is genetic testing to screen for risk of cognitive problems in the future?** | 4.20 (.71)<br>[2 – 5] | 4.48 (.65)<br>[3 – 5] | 4.34 (.69)<br>[2 – 5] |
|  | **% (*n*)** | **% (*n*)** | **% (*n*)** |
| (1) Completely Unacceptable | 0% (0) | 0% (0) | 0% (0) |
| (2) Unacceptable | 4% (1) | 0% (0) | 2% (1) |
| (3) Unsure | 4% (1) | 8% (2) | 6% (3) |
| (4) Acceptable | 60% (15) | 36% (9) | 48% (24) |
| (5) Completely acceptable | 32% (8) | 56% (14) | 44% (22) |
| **Do you have any concerns about genetic testing?** | **% (*n*)** | **% (*n*)** | **% (*n*)** |
| No Concerns | 88% (22) | 84% (21) | 86% (43) |
| Reported Concerns | 16% (4) | 16% (4) | 16% (8) |
| **Blood Draw for Genetic Testing** | **% (*n*)** | **% (*n*)** | **% (*n*)** |
| Completed | 100% (25) | 96% (24) | 98% (49) |
| Not Completed | 0% (0) | 4% (1) | 2% (1) |

Responses to acceptability rated item, percent of participants who reported concerns about genetic testing, and rates of blood draw completion by HIV status group and for the total sample (*N* = 50).

### Concerns

Regarding concerns about genetic testing, eight participants (16%; 8/50) reported at least some concern related to genetic testing. Identical proportions of PHIV (16%; 4/25) and control (16%; 4/25) participants reported concerns. Seven of the eight participants who reported concerns (87.5%) were 18-24 years old and 63.5% identified as female. Comparing participants who reported concerns and participants who did not report concerns, significant differences were found regarding education, *t* (48) = -2.65, *p* = .01, *d* = -1.02, with participants who reported concerns having more years of education (*M* = 13.75 education years, *SD* = 1.58) than participants who did not report concerns (*M* = 10.95 education years, *SD* = 2.89). Furthermore, reporting concerns was significantly associated with post-secondary education, *X*^2^ (1, *N* = 50) = 8.79, *p* = .003, with all eight participants who reported concerns having attended/completed vocational school or college/university. No other demographic associations or differences were found between participants who reported concerns and peers. Participants’ demographic data by concern group are presented in Table 2.

**Table 2.** Participant Demographics by Reported Concerns about Genetic Testing.

|  | Reported Concerns (*n* = 8) | No Concerns (*n* = 42) |
| --- | --- | --- |
|  | % (*n*) | % (*n*) |
| **Age (years)** | *M* = 20.63 (*SD* = 2.45) | *M* = 18.88 (*SD* = 3.15) |
| 13-17 years | 12.5% (1) | 33.3% (14) |
| 18-24 years | 87.5% (7) | 66.7% (28) |
| **Gender Identity** |  |  |
| Female | 62.5% (5) | 50.0% (21) |
| Male | 25.0% (2) | 50.0% (21) |
| Transgender<sup>a</sup>. | 12.5% (1) | 0.0% (0) |
| **HIV Status** |  |  |
| PHIV | 50.0% (4) | 50.0% (21) |
| HIV-negative | 50.0% (4) | 50.0% (21) |
| **Student/Work Status** |  |  |
| Student | 62.5% (5) | 66.7% (28) |
| Employed | 37.5% (3) | 33.3% (14) |
| **Education (years)\*** | *M* = 13.75 (*SD* = 1.58) | *M* = 10.95 (*SD* = 2.89) |
| Vocational School/<br>College/University* | 100.0% (8) | 42.9% (18) |

Demographic data for participants who reported concerns about genetic testing compared to participants who did not report concerns.
a. One participant self-identified as transgender.
\*Significant difference between groups.

Among the eight participants reporting concerns, one participant reported minimal, non-specific concern (“I have only minimal concern”), whereas seven participants reported at least one specific concern. Specific concerns fell within two overarching categories: A) concerns about the risk of genetic/personal data being shared (*n* = 4); B) concerns about the potential for blood samples and genetic data being collected or used in an unethical, illegal, or unknown way (*n* = 4). Three of the four participants who reported concerns about the risk of genetic data being shared with others (i.e., category A) were PHIV participants. Conversely, all four participants who reported concerns about the potential for genetic data being misused (i.e., category B) were control participants. Table 3 presents participants’ reported concerns about genetic testing.

**Table 3.** Participants’ Reported Concerns about Genetic Testing.

| Concern Theme | Reported Concern |
| --- | --- |
| A. Concerns about the risks of genetic/personal data being disclosed or shared ($n = 4$). | *I am concerned about the risk of data disclosure.* (PHIV, female, 18-24 years old) |
|  | *I am worried about the risk of disclosing my genetic characteristics to others.* (PHIV, female, 12-17 years old) |
|  | *I am concerned about others knowing my personal information.* (PHIV, female, 18-24 years old) |
|  | *I am worried about potential data leakage...* (HIV-negative, male, 18-24 years old)<sup>a</sup> |
| B. Concerns about blood samples or genetic data being collected or used in an unethical, illegal, or unknown way ($n = 4$). | *I am slightly concerned if my blood or the testing results will be used in other different types of study...* (HIV-negative, female, 18-24 years old) |
|  | *I have no concern regarding how the genetic study will be conducted. However, I am worried that the genetic information could be used illegally and unethically.* (HIV-negative, male, 18-24 years old) |
|  | *Consent must be obtained prior to every genetic research. People need to be explained the study's objectives clearly to avoid misunderstanding and misuse of data i.e., against the law...* (HIV-negative, female, 18-24 years old) |
|  | *...it [data] might be used in an illegal or unethical way.* (HIV-negative, male, 18-24 years old)<sup>a</sup> |
| C. Minimal, non-specific concern ($n = 1$). | *I have only minimal concern.* (PHIV, transgender; 18-24 years old) |

Eight participants reported concerns about genetic testing. Participants' responses were recorded in Thai and translated into English by study staff. Participants' HIV status, self-identified gender, and age group are presented in parentheses.
a. One participant reported two specific concerns.

### Feasibility

Of the 50 participants recruited to participate in this pilot study, 49 participants (98%) agreed to genetic testing and provided blood samples for this purpose. Notably, all 25 PHIV participants (100%) agreed to genetic testing. Only one control participant declined to genetic testing. When asked their reason for declining, this participant reported:

> *I have just undergone a blood draw for my health check-up recently. I do not want to be hurt, and I am also concerned about my genetic information being shared out*.
>
> (HIV-negative, male, 18-24 years old)

All 49 of the collected blood samples were successfully stored on-site and cold-chained shipped to the US for genetic testing. All collected blood samples (100%) arrived intact and were successfully processed, demonstrating the feasibility of future larger studies of genetic testing in this population.

## Discussion

This pilot study aimed to evaluate the acceptability and feasibility of genetic testing among adolescents and young adults with PHIV and demographically similar HIV-negative peers in Thailand. Findings suggest that genetic testing was both highly acceptable and feasible in our pilot sample, with results exceeding all a priori benchmarks. Among our Thai youth participants, 92% rated genetic testing as either “acceptable” or “completely acceptable,” and 98% of participants agreed to genetic testing and provided blood samples for this purpose. The high acceptability and uptake of genetic testing in the current study aligns with previous findings of generally favorable attitudes towards genetic testing among adults from LMICs.^14,15^ Additionally, all collected blood samples were successfully stored onsite, shipped from Thailand to the US, and processed at a US-based laboratory, suggesting genetic research may be feasible even in settings with limited resources to conduct in-country genetic testing. Overall, findings offer support for future research studies using genetic testing to investigate the genetic underpinnings of HIV-NCI among young people living with PHIV in Thailand.

To further inform planning future genetic studies within this population, we also assessed participants’ concerns about genetic testing. Only eight participants (16%), four PHIV and four control participants, reported concerns about genetic testing. Characterizing theses eight participants, seven (87.5%) were 18-24 years old and all eight received at least some post-secondary education (i.e., vocational school or college/university). Compared to peers, participants who reported concerns had significantly more years of education. Regarding the specific concerns reported, four participants reported concerns about genetic/personal information being shared with others, and four participants reported concerns about the potential for using genetic data in an unethical, illegal, or unknown way. Previous studies among adults from LMICs have also found concerns about confidentiality are common, suggesting an understanding of genetic information as particularly sensitive even among adults with limited knowledge about genetic testing.^14^ Three of the four participants (75%) who reported concerns about genetic/personal data being shared were PHIV participants, whereas all four participants who reported concerns about the potential for misusing genetic data were control participants. Notably, seven of the eight participants who reported concerns also agreed to genetic testing and provided blood samples. This finding may be related to reported concerns not rising to the level to deter participation. Alternatively, it is possible that some participants may agree to genetic testing even though they have concerns about or feel uncomfortable with genetic testing. Nevertheless, findings highlight the need for future genetic studies among young people with HIV in Thailand and other LMICs to explicitly address possible concerns about genetic testing with participants during the informed consent process. Greater effort to actively elicit and address related concerns may be needed with adolescent and young adult participants. Studies of young adults (18-24 years) with post-secondary education or that recruit from collegiate settings may particularly benefit from discussing with participants the specific safeguards taken by the research team to minimize the risk of data disclosure and clearly explaining how collected samples and genetic data will and will not be used.

This study has several limitations worth considering. Although the sample size was appropriate to achieve the aims of this pilot study, the limited sample size necessitates consideration. It is possible that the high acceptability of genetic testing found in our pilot sample may differ from that of the larger population of Thai adolescents and young adults living with HIV. Future genetic studies among Thai adolescents and young adults with larger sample sizes are warranted. Additionally, the acceptability items were specific to genetic testing to assess risk of future neurocognitive problems. It is unclear if genetic testing for other specific purposes would also be equally acceptable among these young people, and future research is needed to clarify this. Lastly, concerns about genetic testing were documented via write-in text response, which did not allow follow-up inquires to clarify or expand participants’ initial responses. Likewise, we did not assess participants’ perceptions of or knowledge about genetic testing, genetic risk, or neurocognitive problems. Future qualitative studies are needed to capture related perspectives and attitudes among these youth more fully. Despite these limitations, this pilot study meaningfully contributes to the literature in offering support for the acceptability and feasibility of genetic testing in research among adolescents and young adults living with HIV in Thailand and in offering insights that can be used to inform the planning of future related studies.

## Conclusions

This pilot study is among the first to support the acceptability and feasibility of research investigating the genetic underpinnings of HIV-NCI among young people living with HIV in an LMIC. Moreover, we assessed acceptability both in terms of participant ratings and actual blood draws completion rates for genetic testing, rather than relying solely on participants’ self-reported willingness to complete genetic testing. Among this sample of Thai adolescents and young adults living with and without HIV, genetic testing to assess risk of future neurocognitive problems was found to be highly acceptable. This study also found genetic testing to be highly feasible in a model where specimens were collected in the LMIC and shipped to the US for processing, using a widely available commercial shipping service. Models to scale up genetic testing in research and related clinical services should be explored further in young people living with HIV in resource-limited settings and other distinct cultural contexts.

## Data Availability

All data produced in the present study are available upon reasonable request to the authors.

## Acknowledgments

We would like to acknowledge all our participants and the research team at the Research Institute for Health Sciences, Chiang Mai University.

## Authors’ Contributions

A.F.S led the study conceptualization and design, data analyzes, writing the original draft, and revising the manuscript. C.M.F. and M.A. assisted with writing the original draft and revising the manuscript. L.A. oversaw data collection, supervised the study team in Thailand, and reviewed and revised the manuscript draft. D.K. provided mentorship and reviewed and revised the manuscript draft. R.N.R. led the parent study, provided mentorship, contributed to the study design and analytic plan, and reviewed and revised the manuscript draft.

## Author Declaration

This work was conducted under ethical approval from the IRB at the New York State Psychiatric Institute and received a Certification of Ethical Clearance from the Human Experimentation Committee at the Research Institute for Health Sciences, Chiang Mai University.

## References

1. World Health Organization (WHO). Maternal, Newborn, Child and Adolescent Health. 2021. Available from: https://www.who.int/maternal_child_adolescent/topics/adolescence/hiv/en/, accessed January 25, 2022.

2. Laughton B, Cornell M, Boivin M, et al. Neurodevelopment in perinatally HIV-infected children: A concern for adolescence. J Int AIDS Soc 2013;16(1):18603; doi: 10.7448/IAS.16.1.18603.

3. Garvie PA, Zeldow B, Malee K, et al. Discordance of cognitive and academic achievement outcomes in youth with perinatal HIV exposure. Pediatr Infect Dis J 2014;33(9):e232–e238; doi: 10.1097/INF.0000000000000314.

4. Nachman S, Chernoff M, Williams P, et al. Human immunodeficiency virus disease severity, psychiatric symptoms, and functional outcomes in perinatally infected youth. Arch Pediatr Adolesc Med 2012;166(6):528–535; doi: 10.1001/archpediatrics.2011.1785.

5. Olivier I, Cacabelos R and Naidoo V. Risk factors and pathogenesis of HIV-associated neurocognitive disorder: The role of host genetics. Int J Mol Sci 2018;19(11):3594; doi: 10.3390/ijms19113594.

6. Gill AJ, Garza R, Ambegaokar SS, et al. Heme oxygenase-1 promoter region (GT)n polymorphism associates with increased neuroimmune activation and risk for encephalitis in HIV Infection. J Neuroinflammation 2018;15(1):70; doi: 10.1186/s12974-018-1102-z.

7. Garza R, Gill AJ, Bastien BL, et al. Heme oxygenase-1 promoter (GT)n polymorphism associates with HIV neurocognitive impairment. Neurol - Neuroimmunol Neuroinflammation 2020;7(3):e710; doi: 10.1212/NXI.0000000000000710.

8. Mukerji SS, Locascio JJ, Misra V, et al. Lipid profiles and APOE4 allele impact midlife cognitive decline in HIV-infected men on antiretroviral therapy. Clin Infect Dis 2016;63(8):1130–1139; doi: 10.1093/cid/ciw495.

9. Yang FN, Bronshteyn M, Flowers SA, et al. Low CD4+ cell count nadir exacerbates the impacts of APOE ε4 on functional connectivity and memory in adults with HIV. AIDS 2021;35(5); doi: 10.1097/QAD.0000000000002840.

10. Maltese PE, Poplavskaia E, Malyutkina I, et al. Genetic tests for low-and middle-income countries: A literature review. Genet Mol Res 2017;16(1); doi: 10.4238/gmr16019466.

11. Zhong A, Darren B, Loiseau B, et al. Ethical, social, and cultural issues related to clinical genetic testing and counseling in low-and middle-income countries: A systematic review. Genet Med 2021;23(12):2270–2280; doi: 10.1038/s41436-018-0090-9.

12. Posner J, Biezonski D, Pieper S, et al. Genetic studies of mental illness: Are children being left behind? J Am Acad Child Adolesc Psychiatry 2021;60(6):672–674; doi: 10.1016/j.jaac.2020.12.031.

13. Stewart KFJ, Kokole D, Wesselius A, et al. Factors associated with acceptability, consideration and intention of uptake of direct-to-consumer genetic testing: A survey study. Public Health Genomics 2018;21(1-2):45–52; doi: 10.1159/000492960.

14. Meilleur KG, Coulibaly S, Traoré M, et al. Genetic testing and counseling for hereditary neurological diseases in Mali. J Community Genet 2011;2(1):33–42; doi: 10.1007/s12687-011-0038-0.

15. Basson F, Futter MJ, Greenberg J. Qualitative research methodology in the exploration of patients’ perceptions of participating in a genetic research program. Ophthalmic Genet 2007;28(3):143–149; doi: 10.1080/13816810701356627.

